# Cardiologists’ perspectives on sociocultural and structural factors shaping cardiovascular genetic testing

**DOI:** 10.64898/2026.06.22.26356233

**Authors:** Heather M. Ramey, Jazmine Gabriel, Ana Morales, Katrina Romagnoli, Marc S. Williams

## Abstract

**Introduction:** Genetic testing is increasingly central to the diagnosis and management of cardiovascular genetic conditions. However, use and follow-through vary across patient populations. Examining clinician perspectives on sociocultural and structural factors influencing testing is important for understanding these differences and informing public health genomics research and implementation efforts.

**Methods:** We conducted semi-structured interviews with 15 cardiologists from health systems across the United States who have integrated cardiogenetics in their practice. Interviews explored experiences diagnosing cardiovascular genetic conditions among patients from underrepresented backgrounds, as well as approaches to incorporating social and contextual information into care. Data were coded thematically and analyzed using a framework analysis guided by the Health Equity Implementation Framework and Social Determinants of Health domains.

**Results:** Clinicians described multi-level factors shaping genetic testing practices, including patient-provider interactions, clinical workflows, health system infrastructure, and broader policy contexts. Key themes included challenges communicating complex genetic information across language and literacy differences; patient trust shaped by prior healthcare experiences; fragmented insurance coverage separating genetic testing from genetic counseling; and challenges interpreting variants of uncertain significance, particularly for populations underrepresented in genomic reference databases. Clinicians also described adaptive strategies—such as interdisciplinary collaboration, telehealth, and patient assistance programs—that supported testing in some settings but were often inconsistent or resource-dependent.

**Conclusion:** Among cardiologists using genetic testing, system-level and sociocultural factors shape the feasibility and downstream use of cardiovascular genetic testing. Findings highlight considerations for public health-informed genomic infrastructure that accounts for social context, supports communication, and reduces reliance on individual clinician workarounds, with implications for clinical decision support and related public health genomics initiatives.

## Introduction

Cardiovascular conditions account for approximately one-third of all deaths worldwide, making them the leading cause of global mortality [1]. The global burden is expected to rise due to aging populations [2], making this a significant public health concern and underscoring the importance of early detection and effective management. While many cardiovascular conditions can be identified and prevented by addressing modifiable risk factors, family history, monogenic disease, and genetic variation identified by polygenic risk scores play a critical role in specific conditions [3], including cardiomyopathies, lipid disorders, and hereditary amyloidosis. Recognition of genetic risk—coupled with enhanced access to genetic testing—can shorten diagnostic timelines and inform treatment, family-based testing, and risk management.

Underrepresentation of many populations in genomic reference databases [4] limits generalizability of genetic findings and risks widening disparities in cardiovascular genomics. Beyond biology, factors outside of the clinical encounter, including social context, structural barriers, and resource constraints, shape who is offered, completes, and ultimately benefits from genetic testing. These challenges disproportionately affect historically marginalized and under-resourced communities [5–7]. Social determinants of health (SDOH) provide a framework for understanding how these upstream factors—including economic stability, education access, health care access, neighborhood and built environments, and social and community context—affect patients’ diagnostic journeys and downstream care.

SDOH are recognized by the American Heart Association (AHA) as key drivers of cardiovascular risk and outcomes [8] and are integral in achieving the goals in Healthy People 2030 and the United Nations’ Sustainable Development Goals [9–10]. Positioning SDOH within cardiovascular genetics care highlights how social context and structural conditions intersect with genetic risk to influence timeliness and accessibility of care. These considerations highlight the need to understand how clinicians perceive and navigate the sociocultural and contextual factors that shape genetic testing in real-world cardiovascular practice, as clinicians have a central role in determining when testing i s offered, how risk is communicated, and which patients are able to access genetic services. This work extends a larger NIH-funded project focused on the pre-implementation of a clinical decision support (CDS) tool designed to lower barriers to identifying individuals who may benefit from genetic testing [11]. The goal of this study was to examine how sociocultural factors influence the completion of cardiovascular genetic testing from the clinicians’ perspective. Clinicians were asked about 1) their experiences diagnosing patients from underrepresented communities and 2) how SDOH were integrated into patient care. By centering clinicians’ experiences across diverse settings, this work aims to inform equitable CDS design that accounts for contextual factors and SDOH.

## Methods

### STUDY DESIGN

Semi-structured qualitative interviews were employed and analyzed using a framework-informed approach. The Health Equity Implementation Framework (HEIF) [12] and the SDOH [10] informed development of the interview guide and structured the analytic codebook, enabling multilevel examination of access, implementation, and equity in genetic testing. These frameworks informed topic areas and were applied most explicitly during coding and analysis. The interview guide was adapted from guides developed in previous work by the study team [11] and was intentionally open-ended; HEIF and SDOH domains informed topic areas rather than serving as a prespecified structure. Ethical approval and consent procedures are described in the Statement of Ethics.

### SETTING

The prime study site is located in the eastern United States (i.e., central and northeast Pennsylvania), where individuals have relatively short distances to cardiology providers [13] and the patient population is predominantly rural, low-income, and white, with low out-migration [14]. To reduce selection bias and enhance transferability, recruitment extended beyond the local geographical area to include clinicians from the grant sites (i.e., Geisinger, Intermountain Healthcare, University of Utah) and external institutions (Tab.1).

### PARTICIPANTS

Participants were purposively sampled to include cardiologists practicing in the United States with experience diagnosing cardiovascular genetic conditions [15,16]. Potential participants were identified by team members and recruited via direct email, with additional participants recruited through snowball sampling. Interviews were scheduled for 45 minutes with one of two interviewers (H.M.R. & J.G.) between February and August 2025. Sampling continued until thematic saturation was achieved [17]. All participants provided informed consent.

### DATA COLLECTION AND INTERVIEW GUIDE

The interview guide from our previous work was revised and expanded to address cardiovascular genetics and incorporate sociocultural context; two open-ended questions were added to elicit perspectives of sociocultural themes (Appendix I). The HEIF and SDOH domains guided both the interview domains and construction of a structured codebook (Appendix II).

### DATA ANALYSIS

Interviews were recorded using Microsoft Teams, then transcribed by a professional medical transcriptionist. Analysis followed a matrix-based Framework Method [18,19], guided by HEIF and SDOH frameworks (H.M.R.). Transcripts were imported into Atlas-ti to support manual deductive coding at the domain level, with additional inductive codes applied within domains to capture content not covered by the initial framework. Coded excerpts were then charted into a Microsoft Excel matrix with framework domains as column headers.

Within each HEIF domain, an inductive thematic analysis was conducted to identify, name, and refine themes based on patterns across excerpts, while remaining grounded in the overarching deductive framework. Codes were organized into themes, which were reviewed iteratively by the qualitative team (J.G. & A.M.) to ensure consensus, clarity, and alignment with study aims, with refinements made during matrix charting and theme development. Mention counts were tracked and defined as the total number of times a theme was referenced across all interviews (i.e., a single participant could contribute more than one mention count if the theme arose multiple times within their interview). Following theme development, coded excerpts were mapped to SDOH domains, and these mappings were summarized across themes and HEIF levels to characterize the distribution of sociocultural and structural influences (Fig. 1).

**Fig. 1.**
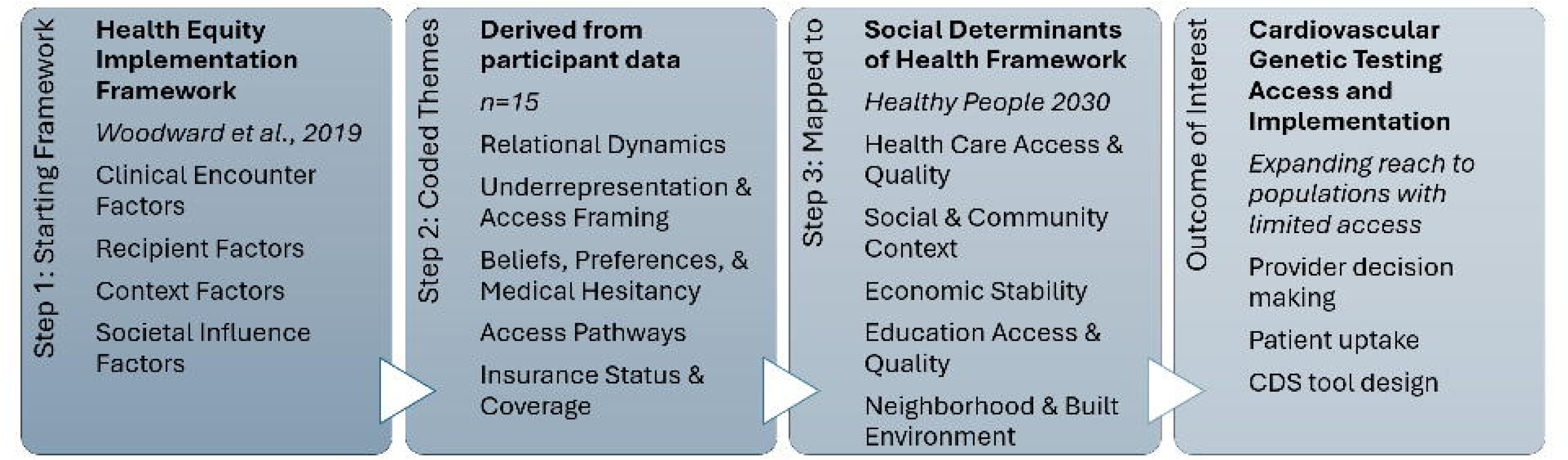
Analytic framework from HEIF domains to emergent themes to social determinants of health. Analytic framework depicting the three-step process used to examine sociocultural factors in cardiovascular genetic care. Health Equity Implementation (HEIF) domains served as the coding structure for participant transcripts (n=15). Emergent themes were identified iteratively and subsequently mapped to Social Determinant of Health (SDOH) domains (Healthy People 2030) to inform analysis. Alt text: Relationship between framework-informed analysis and study findings. Health Equity Implementation Framework domains were used to structure coding of interview data, followed by inductive identification of themes such as relational dynamics, access framing, and medical hesitancy. Themes were then mapped to social determinants of health to support interpretation of sociocultural and structural factors influencing cardiovascular genetic testing.

We used the Standards for Reporting Qualitative Research (SRQR) guidance to inform manuscript development, and the accompanying SRQR checklist was used during final editing and is provided as a supplementary file [20,21].

Microsoft Copilot was used to support manuscript editing and review during preparation of this manuscript. The accuracy of AI-assisted outputs was verified through review. All analytic decisions, including coding, theme development, selection, and interpretation, were made by the primary researcher.

### REFLEXIVITY

Interviews were conducted by researchers trained in qualitative methods and with experience in genomic health research (H. M.R.: public health; J.G.: genetic counseling). H.M.R. led coding, matrix development, and initial theme identification. To enhance analytic rigor and transparency, we grounded the codebook in HEIF and SDOH, maintained analytic memos, and used iterative team review to refine matrices and theme names; J.G. & A.M. contributed to theme review and consensus.

## Results

### PARTICIPANTS

Fifteen cardiology clinicians participated (C001 through C016), representing varied subspecialties and practice settings; C010 is excluded from analysis as the interview concluded prior to the administration of the sociocultural domain questions (Tab. 1).

**Tab. 1.** Participant demographics and clinical characteristics. Characteristics of cardiologist participants (n=15) including provider type, subspecialization, institutional affiliation, years in practice, and populations served.

|  | ***n=15*** |
| --- | --- |
| **Provider type** |  |
| Physician | 14 |
| Physician Assistant | 1 |
| **Sub-specialization** |  |
| Adult practice | 13 |
| Advanced heart failure and transplantation | 7 |
| Imaging (invasive and non-invasive) | 5 |
| General cardiology | 3 |
| Pediatric practice | 2 |
| **Expertise** |  |
| Research experience | 12 |
| Multidisciplinary clinic experience | 6 |
| Lipidology | 2 |
| Sports exercise | 1 |
| Clinical informatics | 1 |
| **Affiliation** |  |
| Cleveland Clinic | 2 |
| Geisinger Health System | 2 |
| Intermountain Health | 2 |
| Stanford Medicine | 2 |
| Morehouse School of Medicine | 1 |
| Oregon Health & Science University | 1 |
| Texas Children's Hospital | 1 |
| University of California San Diego | 1 |
| University of Florida | 1 |
| University of Texas Health & Science Houston | 1 |
| University of Utah | 1 |
| **Years in practice** |  |
| <5 | 2 |
| 5-10 | 3 |
| 11-20 | 5 |
| 20+ | 5 |
| **Populations Served \*** |  |
| Hispanic | 8 |
| Rural/Underserved | 7 |
| Black/African American | 6 |
| Immigrant | 2 |
| South Pacific Islander | 2 |
| Asian | 1 |
| Latino | 1 |
| Men/Women | 1 |
| Native American | 1 |
| Non-English Speaking | 1 |
| Non-Citizens/Undocumented | 1 |
| Patients Living in Poverty | 1 |
| Portuguese | 1 |
| Persons with Substance Use Disorder | 1 |
| South American | 1 |
| Vietnamese | 1 |

*\* Populations served reflect categories mentioned by participants in response to open-ended questions about caring for patients from underrepresented populations. Responses were provided as free text rather than selected from a predefined list. Participants could reference more than one population, and not all participants commented on the populations they served. Counts are not intended to represent comprehensive clinic demographics.*

### PATIENT POPULATIONS

Participants described caring for patients from diverse sociocultural and structural backgrounds (Tab. 1). Based on clinical experience, some groups were associated with specific cardiovascular genetic conditions (e.g., atypical Marfan syndrome presentations among Hispanic patients, heart failure and hereditary amyloidosis among Black patients, transthyretin-associated amyloidosis among Portuguese patients, familial hypercholesterolemia among Lebanese patients, and cardiomyopathy in the context of substance use disorders).

### THEMATIC ANALYSIS

Findings were organized using the four HEIF analytic layers: Clinical Encounter (Patient-Provider Interaction), Recipients (Provider, Patient), Context (Local, Organizational, Healthcare System), and Societal Influence (Economics, Physical Structures, Sociopolitical Forces). While themes are presented within discrete HEIF domains for analytic clarity, some interaction-level experiences—such as language barriers—are examined across multiple interaction-level themes (e.g., Language & Communication and Relational Dynamics) to reflect both communication challenges and effects on relational processes, including trust, rapport, and patient engagement. Numbers in parentheses indicate mention count, defined as the number of times a theme was referenced across interviews. A complete list of domains, themes with mention count, examples, mapping to SDOH categories, and exemplary quotes can be found in Appendix III; a summarized version is presented in Table 2.

**Tab. 2.**
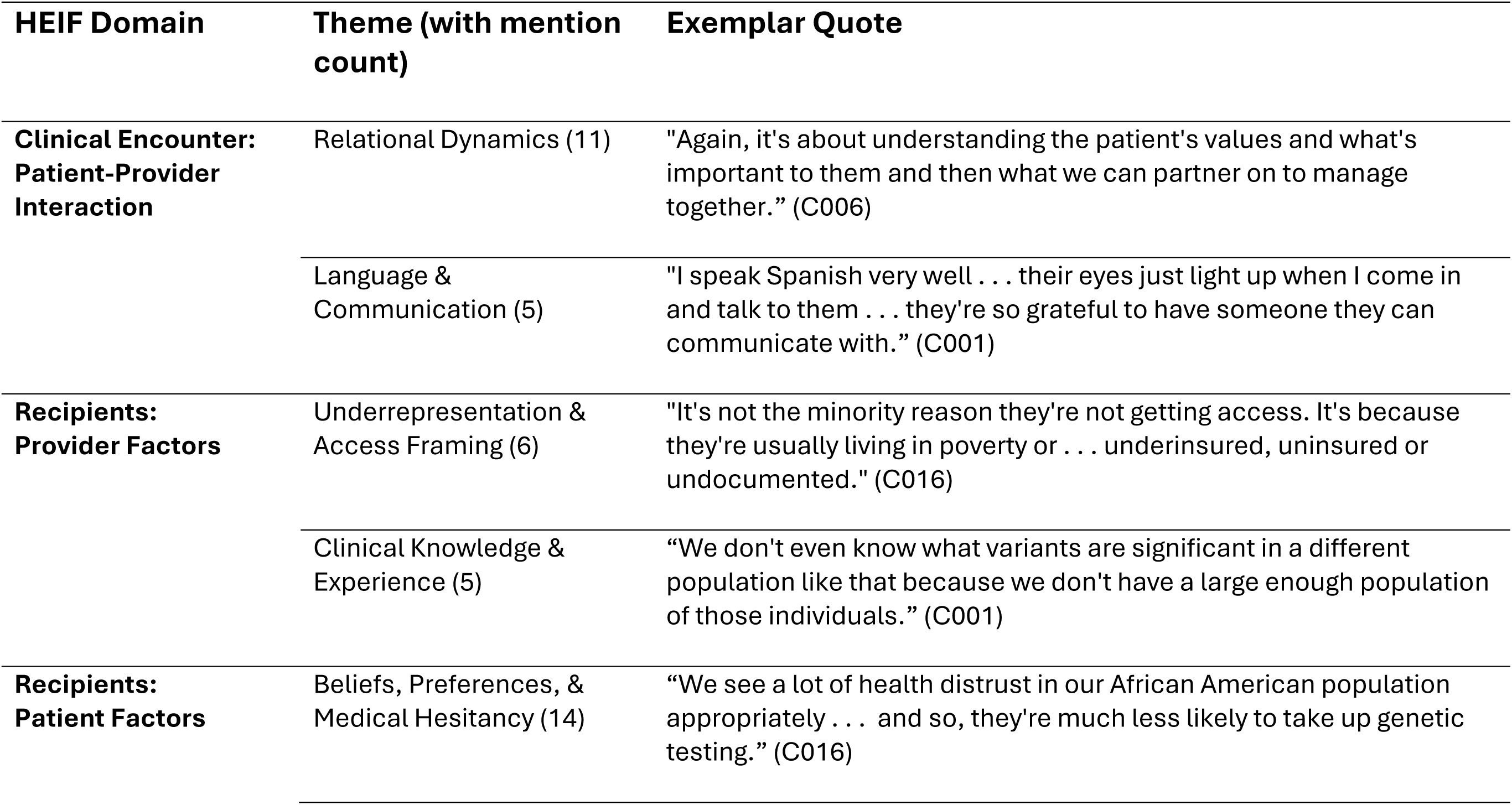
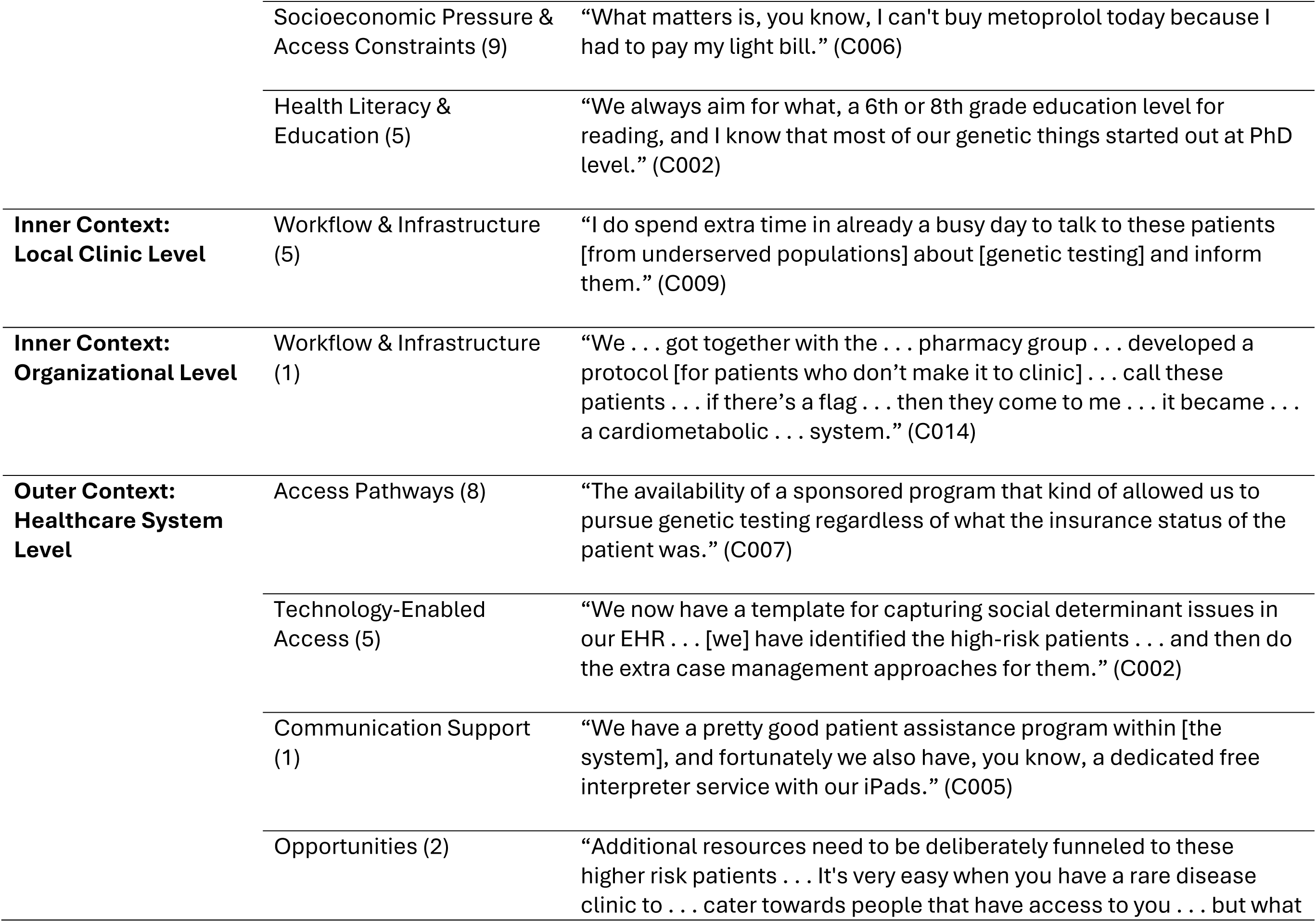
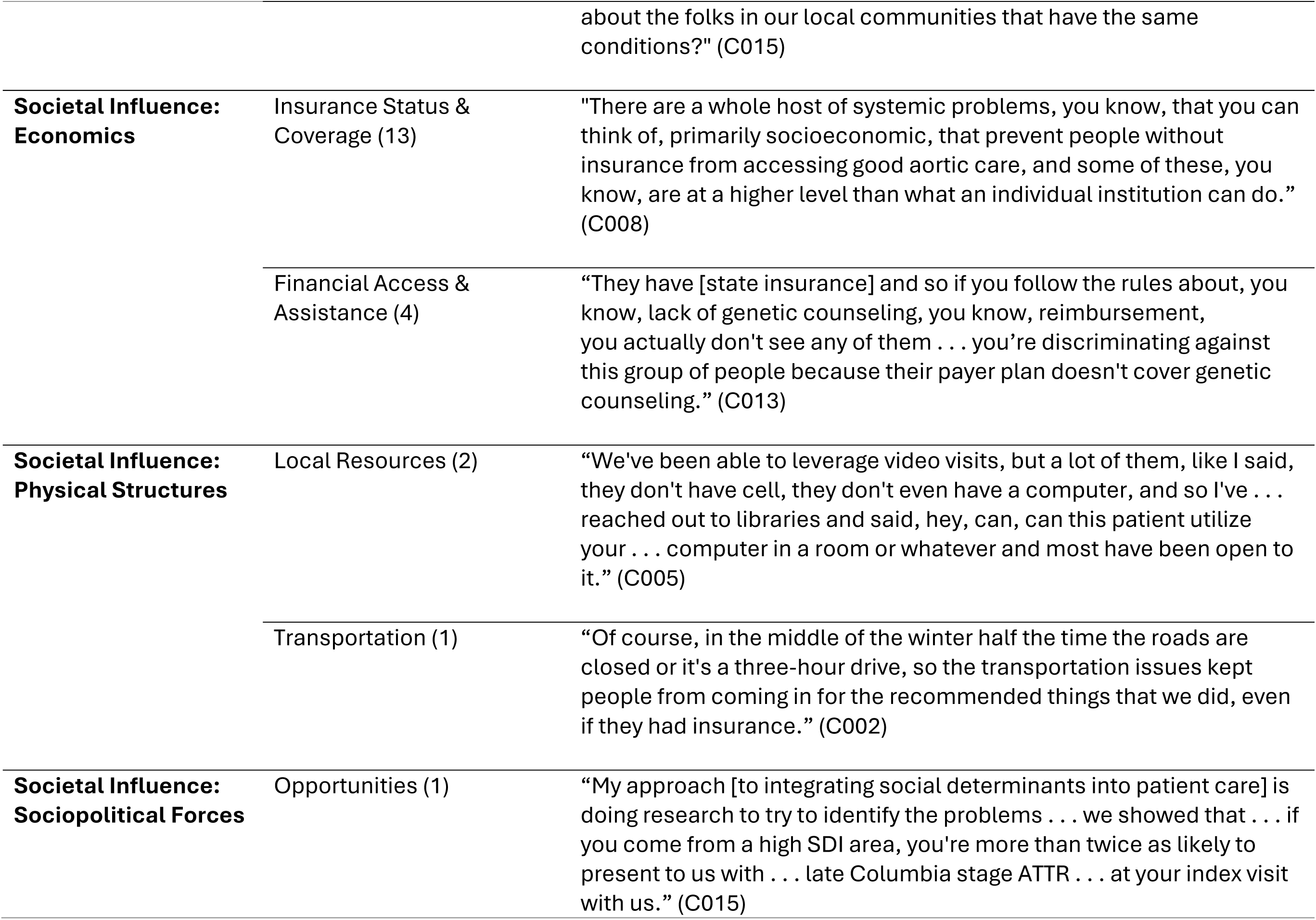
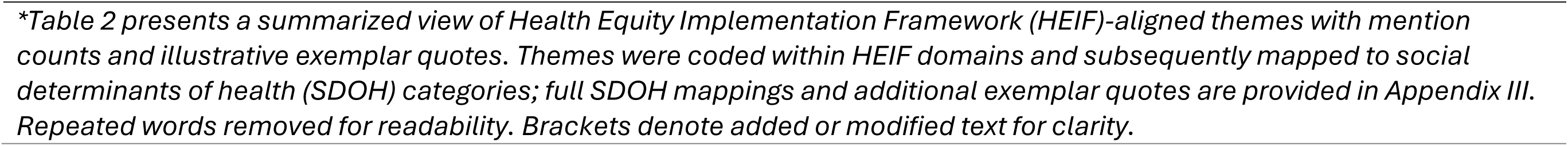
HEIF-aligned themes and exemplar quotes. *. Summary of themes identified through framework analysis, organized by Health Equity Implementation Framework (HEIF) domain, with mapped Social Determinants of Health (SDOH) categories and supporting participant quotes. Full theme list presented in Appendix III.

#### Patient-Provider Interaction Factors

Two themes were identified: Relational Dynamics and Language & Communication.

##### Relational Dynamics (11)

Clinicians described how time pressure and limited language concordance contributed to patients feeling rushed and being less likely to pursue care, constraining rapport-building during visits (C013). Conversely, listening and rapport-building increased receptiveness to testing recommendations (C006, C014, C015). One clinician emphasized that “establishing that relationship during the first visit” supported trust to introduce the subject of genetic testing (C013). Limited peer support was linked to a shortage of genetic counselors from underserved communities, reducing opportunities for culturally concordant relationship-building (C004). Family presence during visits supported comprehension and planning for cascade testing (C015). Culture-related beliefs shaped perceptions of the medical system (C003) and engagement of relatives (C003, C009), with some patients not being comfortable contacting clinicians, contributing to drop-off (C005).

##### Language & Communication (5)

Limited English proficiency impeded navigation and communication, which was associated with perceived discrimination (C013). Even with interpreters, clinicians described challenges in conveying depth and nuance (C009, C013). In contrast, language concordance facilitated engagement; clinicians noted translator impact (C001) and positive patient responses when provider spoke their native language (C001).

#### Provider Factors

Two themes were identified: Underrepresentation & Access Framing and Clinical Knowledge and Experience.

##### Underrepresentation & Access Framing (6)

Participants primarily framed underrepresentation through socioeconomic access rather than race or ethnicity; emphasizing that insurance status and financial strain more strongly impacted those who were able to access specialty cardiovascular genetic care. Examples included challenges rooted in “poverty or uninsured, underinsured . . . or undocumented” status (C016), and for patients with Medicaid (C015). It was noted access challenges occurred upstream of the clinical encounter, for example, “who ends up coming to our clinic . . . that’s harder, right?” (C012). Moreover, testing decisions were based on clinical criteria rather than ethnicity (C014) and patient concerns were described as “cut[ting] across racial barriers,” with privacy emerging as a prominent concern that was perceived as more influential than race or ethnicity in shaping patients’ testing decisions (C007). In contrast to adult practices, in pediatric settings, “one of the benefits” is comprehensive insurance coverage as it enables providers to care for patients the same way regardless of background (C011).

##### Clinical Knowledge & Experience (5)

Participants described uncertainty about variant interpretation in underrepresented populations (C001), variable awareness of syndromic aortopathy manifestations by racial and ethnic backgrounds (C008), and difficulty parsing behavioral causes from genetic contributions in cardiomyopathy (C009). Some were unsure how social or cultural factors impact patients’ testing decisions (C012). Less exposure to underrepresented groups limited opportunities to recognize varied presentations (C003).

#### Patient Factors

Three themes were identified: Beliefs, Preferences, & Medical Hesitancy, Socioeconomic Pressure & Access Constraints, and Health Literacy & Education.

##### Beliefs, Preferences, & Medical Hesitancy (14)

Clinicians described patients declining invasive therapies (e.g. “I don’t want to be cut on” [C006]), expressing privacy concerns including third-party testing (C007), or being hesitant to learn genetic information (C001, C016). Others reported patients who were brought in by family who were otherwise disengaged in healthcare (C001), while some patients demonstrated trust in healthcare and a “thirst for knowledge” related to family health (C012). Family dispersion or estrangement due to geographical separation complicated outreach (C003, C013), while bringing family to visits supported comprehension and planning (C003).

Participants perceived broad distrust of healthcare among Black and African American and rural communities, which was described as being compounded by the relational and familial implications of cardiovascular genetic care, particularly concerns related to family communication, pride, and disclosure of potential risk to relatives (C001, C004, C005, C011, C016).

##### Socioeconomic Pressure & Access Constraints (9)

Competing demands were frequently described as displacing genetics-related care, particularly among patients experiencing socioeconomic constraints. Co-morbid conditions (e.g., diabetes, substance use disorder) took precedence (C009). Clinicians described trade-offs between medication adherence and basic needs (C006), inflexible work schedules (C012, C013), limited connectivity across large rural catchments (C005), financial and insurance limitations (C003, C007), and transportation challenges for multi-visit plans (C012). Federal and state assistance programs were cited as facilitators (C005).

##### Health Literacy & Education (5)

The technical complexity of genomics (C002) posed recurring barriers to informed consent and meaningful engagement with results (C002, C003, C009, C012).

#### Context Factors

Five themes were identified across context levels: Workflow & Infrastructure at the Local Clinic Level and the Organizational Level; Access Pathways, Technology-Enabled Access, Communication Support, and Opportuni ties at the Healthcare System Level.

##### Workflow & Infrastructure (Clinic) (5)

Clinicians described time pressure in clinic schedules as a barrier to thorough consent discussions, particularly when language barriers increased the time required for explanation and interpretation (C009). Clinicians also flagged privacy considerations tied to third-party vendor testing platforms (C007). Adaptations included clinic-level workflow changes to mitigate payer and system constraints, such as coordinating clinician and genetic counseling evaluations on the same day (C013) and leveraging transition programs to build health literacy in adolescence (C016).

##### Workflow & Infrastructure (Organizational) (1)

One participant cited cross-departmental adaptations that preserved patient safety when access was delayed (e.g., medication continuity) (C014).

##### Access Pathways (8)

Clinicians described sponsored or patient assistance programs (C005, C007), social work support (C016), parking passes (C011), partnerships with community organizations (C001, C006), and inclusivity and outreach efforts (C001, C011) as facilitators.

##### Technology-Enabled Access (5)

Telehealth was described as a facilitator for rural patients, enabling access to specialty care while mitigating transportation barriers (C014), as was electronic health record (EHR)-based screening to identify eligible patients (C008) or social needs that could impede testing (C002, C006).

##### Communication Support (1)

Free interpreter services were described as facilitators (C005). Opportunities (2). Clinicians described potential for targeting additional resources to “higher-risk patients” (C015) and aligning more closely with local community health initiatives (C006).

#### Societal Influence Factors

Five themes were identified: Insurance Status & Coverage and Financial Access & Assistance at the Economics Level; Local Resources and Transportation at the Physical Structures Level; and Opportunities at the Sociopolitical Forces Level.

##### Insurance Status & Coverage (13)

Clinicians frequently reported that patients either lacked adequate insurance for genetic services, including absence of coverage, underinsurance, or payer limitations that excluded genetic testing or counseling—even when patients were otherwise insured (C007, C008, C011, C012, C013, C015, C016). Coverage-related gatekeeping was described as restricted gene panels despite phenotype (e.g., limiting familial hypercholesterolemia testing to a subset of genes [C014]) and the lack of payer reimbursement for genetic counseling, which clinicians described as effectively excluding patients from services and as discriminatory in practice (C013). In response, some clinicians described clinic-level adaptations to mitigate exclusion, including coordinating genetic counseling and physician evaluations within the same visit so that patients could be scheduled and evaluated despite lack of standalone counseling reimbursement (C013). In contrast, some clinicians noted that once patients reached the specialty clinic or pediatric setting with guaranteed coverage, insurance was less likely to impede genetic testing (C007, C011, C012).

##### Financial Access & Assistance (4)

Clinicians described financial barriers including cost differences between public and private clinical settings (C008), and out-of-pocket expenses that were prohibitive for some patients (e.g., testing costs) (C007). Some patients relied on government-based resources to offset costs, although these supports were not always adequate (C014). One clinician described collaborating with pharmacy teams to identify copay cards to facilitate access to medications for some patients (C011).

##### Local Resources (2) and Transportation (1)

Participants discussed leveraging charitable or public transportation options (C001), library access for telemedicine (C001), and winter road closures (C002) as factors shaping access.

##### Opportunities (1)

One clinician identified opportunities to study delays and develop interventions that empower local clinicians to initiate genetic testing and to streamline referral pathways (C015).

#### Social Determinants of Health

Across HEIF domains, SDOH mapping concentrated in Health Care Access and Quality (Fig 2a), but the dominant domain varied by level (Fig. 2b). Numbers in parentheses reflect the number of distinct themes within each HEIF domain; accompanying excerpt counts reflect the number of coded excerpts contributing to those themes. SDOH categories were assigned at the level of coded excerpts; percentages reflect the distribution of SDOH mappings aggregated across excerpts within each HEIF level rather than counts of themes.

**Fig 2.**
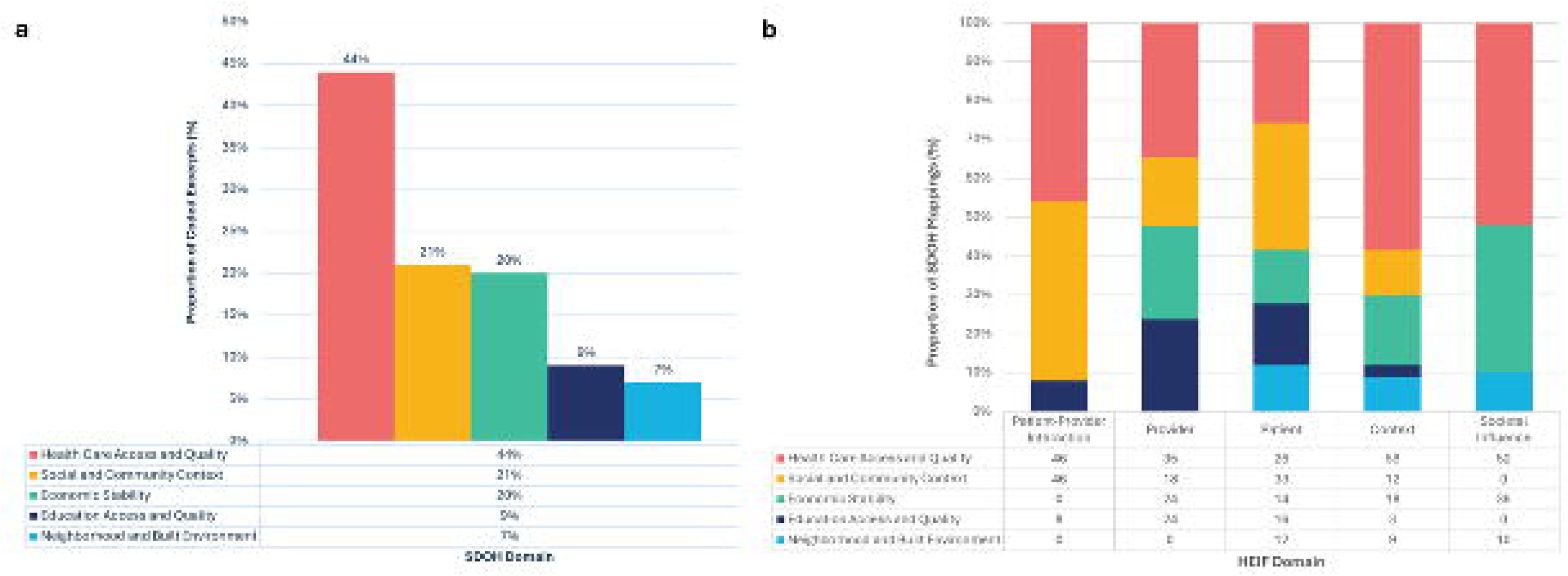
Mapping of social determinants of health across HEIF-aligned analytic themes and levels. Fig. 2. a. Social determinants of health domains mapped across HEIF-aligned themes. Bar graph showing the proportional distribution of Social Determinant of Health (SDOH) domains mapped across Health Equity Implementation Framework (HEIF)–aligned analytic themes. A total of 157 SDOH mapping instances were identified across 98 theme-level analytic instances; coded excerpts could be mapped to more than one SDOH domain. Counts within each SDOH domain were summed and divided by the total number of SDOH mappings to generate percentages. Percentages reflect proportional distribution of SDOH mappings across all coded excerpts rather than population prevalence or the magnitude of patient-level barriers. Fig. 2. b. Distribution of social determinants of health domains across HEIF levels. Stacked bar graph depicting the relative distribution of Social Determinants of Health (SDOH) domains within each Health Equity Implementation Framework (HEIF) domain, including patient–provider interaction, provider, patient, context, and societal influence. Bars represent the proportion of excerpt-level SDOH mappings within each HEIF domain. Percentages illustrate relative analytic distribution of SDOH domains across implementation levels, rather than the absolute frequency of barriers. Alt text: Proportion of Social Determinants of Health (SDOH) categories mapped to coded interview data. Health Care Access and Quality represents forty-four percent of mappings, followed by Social and Community Context at twenty-one percent, and Economic Stability at twenty percent, with smaller contributions from Education Access and Quality and Neighborhood and Built Environment factors. Distribution varies across implementation levels, with Health Care Access prominent across domains and other categories contributing unevenly depending on context.

Patient-provider interaction themes (5), drawing from 16 excerpts, split largely between Health Care Access (46%) and Social and Community Context (46%), with minor contributions from Education Access and Quality (8%).

Provider themes (2), drawing from 11 excerpts, were balanced across Health Care Access (35%), Economic Stability (24%), Education Access (24%), and Social Context (18%).

Patient themes (3), drawing from 28 excerpts, were spread across Social Context (33%), Health Care Access (26%), Education Access (16%), and Economic Stability (14%), and Neighborhood and Built Environment (12%).

Context-level themes (5), drawing from 22 excerpts, were anchored in Health Care Access (59%), with smaller roles for Economic Stability (18%), Social Context (12%), Built Environment (9%), and Education Access (3%).

Societal influence themes (5), drawing from 21 excerpts, were associated heavily with Health Care Access (54%) and Economic Stability (43%), with a lesser role for Built Environment (3%).

## Discussion

Even among cardiologists experienced in genetic testing, clinicians described structural factors—rather than knowledge or motivation—as the primary constraints on equitable genetic testing, including insurance coverage and counseling fragmentation, language and time demands, and result interpretation uncertainty amplified by underrepresentation of individuals not of Northern European ancestry in genomic resources. Clinicians described being able to act only within the limits these structures allow, as reflected in decisions not to offer genetic testing when costs were prohibitive (C014) and adaptations to clinic workflows to accommodate counseling alongside testing (C013). Participants recognized that they were only able to affect a portion of the overall diagnostic journey for their patients. “[G]etting genetic testing covered” is not an issue “once people get into our clinic” (C012), but informed consent is limited as patients find “their payer plan doesn’t cover genetic counseling” (C013). Recent American Heart Association (AHA) guidance emphasizes that cardiovascular risk reflects interactions between genetic susceptibility and social conditions across the life course, highlighting the importance of considering context when translating genomic tools into clinical care [22].

This multi-site sample (n=15) shows that structural friction persists even at the top of the expertise gradient, implying the possibility of larger gaps in general settings. Participants represented a mix of clinical and professional experience, including involvement in clinical guideline development, experience in research, multidisciplinary clinics, or informatics. Relative to cardiologists in general, these participants, many of whom practice in specialty or academic settings, tended to have greater access to genetics experts, higher familiarity with genetics, and more frequent use of genetic testing [23–26]. Despite this level of experience, ongoing burden related to counseling of variants of uncertain significance (VUS) and uncertainty interpreting results for ancestries that are underrepresented in genomic reference databases are prominent across the sample. This parallels challenges described in pediatric cardiovascular genetics, where workflow complexity, variable access to genetics expertise, and inconsistent EHR processes shape genetic testing use [27], and extend to adult care where genomic reference data remain limited [28].

Social determinants have a significant impact on cardiovascular conditions and health outcomes, with population-level evidence demonstrating graded associations between health-related social needs and overall cardiovascular health [29–30]. Consistent with this literature, clinicians in our study described social factors as shaping genetic service delivery across all HEIF domains (summarized in Figure 2), underscoring how social context permeates cardiovascular genetic care. Across these factors, health care access emerged as the most influential across domains, reflecting not patient reluctance but system-level processes that determine who reaches specialty care, which tests and counseling services are reimbursed, and how clinical time is allocated. These constraints shaped not only uptake of genetic testing but whether genetics entered clinical conversation at all. Social and community context and economic stability further conditioned feasibility, as transportation barriers and competing demands related to employment and comorbid conditions frequently displaced genetic discussions. Education access and neighborhood-level factors were less often described as primary barriers by participants, yet they played an important role in determining follow-through and comprehension, influencing logistics and continuity rather than initial intent. Importantly, although these barriers were commonly described as being experienced by patients, they were produced structurally through payer policies, clinic workflows, and institutional capacity, whereas facilitators—such as sponsored testing programs, interpreter services, social work support, and coordinated clinic models—were predominantly system mediated.

While many of the barriers described here are common to cardiovascular care more broadly, they were amplified in the context of genetics, where increased complexity, consent requirements, familial implications, and ancestry-linked uncertainty transform familiar access challenges into qualitatively different implementation problems. Consistent with recent AHA guidance emphasizing how cardiovascular risk reflects interactions between genetic susceptibility and social conditions [22], these findings underscore the importance of social context when translating genomic tools into practice. Limitations in genomic reference resources further amplify these implementation challenges in genetics by complicating interpretation for patients from ancestries that are underrepresented in existing datasets, reinforcing uncertainty around variant pathogenicity and clinical relevance [28,31]. Together, these findings suggest that improving the use of genetic testing will require interventions that address structural access and workflow design—rather than approaches focused solely on provider motivation or knowledge—and align with the American College of Cardiology and American Heart Association (ACC/AHA) Joint Committee on Clinical Data Standards’ emphasis on optimizing outcomes across the lifespan [32].

Findings from this study have implications for downstream implementation efforts, including the design of CDS and related infrastructure. Across all domains, clinicians described challenges that extended beyond individual clinical knowledge gaps, pointing instead to a need for system-level support that assists with interpretation, communication, and navigation of genetic services. For providers, this includes access to contextualized educational resources and support for variant interpretation, particularly tools that assist clinicians in contextualizing variants using available clinical evidence—including phenotypes, family history, and syndromic patterns—and in communicating uncertainty when genomic reference data are limited, especially for populations underrepresented in existing databases. At the patient and interaction levels, clinicians emphasized the importance of communication support that accounts for language, literacy, trust, and family-level considerations. At the system level, participants described the value of mechanisms that surface coverage information, assistance programs, interpreter services, and care coordination resources at the point of care. Collectively, these findings underscore how implementation tools may be most effective when they support existing clinical workflows and reduce reliance on ad hoc clinician workarounds rather than assuming changes in provider motivation or expertise.

## Limitations

This study has several limitations. First, many clinicians practiced in regions with comparatively less diverse patient populations, which may limit observations about racial and ethnic differences and limit transferability to other settings. Second, the sample intentionally skewed towards clinicians with above-average exposure to genetics; appropriate for eliciting domain-specific insights but potentially underestimating barriers faced in clinics with less infrastructure. Third, perspectives from non-physician team members (e.g., genetic counselors, social workers) were not collected and may surface additional themes related to counseling, navigation, and social resource linkage. Finally, the qualitative data reflects reported experiences in specific organizational contexts; findings should be interpreted with attention to local policy and resource environments.

## Conclusion

This study demonstrates that the relevance and impact of sociocultural and structural factors vary across clinical settings and across the lifespan, shaping not only access to genetic testing but how testing conversations unfold in practice. Addressing these factors at the onset of implementation efforts is essential to avoid reinforcing existing access constraints as genomic tools are introduced into routine care. Integrating sociocultural considerations into clinical infrastructure and decision support represents an opportunity to advance responsible and effective cardiovascular genetic care.

## Acknowledgements

The authors would like to thank the individuals who participated in the interviews for this project. We also thank Zachary Salvati and Darren Johnson for their thoughtful feedback and discussion during analysis and dissemination planning. We gratefully acknowledge Carol Gross-Davis and Kati Hinman for mentorship and advising during earlier stages of this work.

## Statement of Ethics

This study was reviewed and approved by Geisinger Institutional Review Board (IRB #2021-0916). Written informed consent was not obtained for participation in this study. Verbal consent was obtained for participation in this study. This protocol was reviewed and approved by Geisinger Institutional Review Board (IRB #2021-0916). All procedures performed in studies involving human participants were in accordance with the ethical standards of the 1964 Helsinki Declaration and its later amendments or comparable ethical standards.

## Author Contributions

H.M.R. led study design, conducted interviews, performed data collection and analysis, and drafted the manuscript. J.G. conducted interviews, contributed to data collection, and critically reviewed the manuscript. M.S.W. provided study oversight and conceptual guidance. A.M. contributed to analytic oversight, conceptual guidance, and theoretical framing. K.R. contributed to conceptual guidance through leadership in related qualitative and human-centered design work. All authors reviewed and approved the final manuscript and agree to be accountable for their contributions.

## Data Availability Statement

The data that support the findings of this study are not publicly available due to privacy reasons and the small sample size, which may increase identifiability. De-identified data may be made available from the corresponding author upon reasonable request for research purposes that are consistent with the original ethics approval and may be subject to a data use agreement.

## Appendix I – Interview Guide

- For this next set of questions, we are curious about any experiences you may have had with diagnosing genetic heart disease in patients from underrepresented populations.

- Clarifying statement: Examples might be those with non-White European ancestries, or those who might have had different experiences with genetic testing than other people for various reasons.
- Could you describe your experience conducting a genetic evaluation for a patient from an underrepresented group who ends up being diagnosed with a cardiovascular genetic condition?
- Can you tell me about your experience with groups in whom diagnosis tends to be more difficult or who may experience better/worse outcomes?
- Are there any challenges related to any areas of the testing process? [Examples: placing an order, interpretation, disclosure]
- What made it (easy/difficult)?
- What is your approach to integrating social determinants of health in the care of your patients? This includes the environmental conditions in which individuals experience life, such as access to quality education and healthcare, economic stability, neighborhood safety, and social and community support.

- How do these factors influence your patients’ preferences about genetic testing?
- If a need is identified, how do you help these patients? [Examples: codes added to the medical record, referrals placed, online/community resources recommended]
- In what ways, if any, do societal factors play a role in making genetic testing easier or difficult in patients from underrepresented groups?
- How would you describe the culture of inclusivity in your clinic or the community you serve?

## Appendix II – Codebook

The codebook served as the foundation for organizing and interpreting the interview data and allowed for both deductive and inductive theme identification. It structured the HEIF domains into clearly defined codes with accompanying definitions and examples. SDOH and relevant clinician demographics were incorporated during analysis to support contextual interpretation of sociocultural and structural factors. As interviews were analyzed, emergent themes were systematically linked to these deductive codes, allowing for a nuanced interpretation grounded in both theory and data. This approach ensured that contextual factors influencing implementation were captured and organized.

| Code group/<br>HEIF domain | Code | Definition | Example |
| --- | --- | --- | --- |
| Clinical encounter | Innovation | Relative fit and degree of fit within existing practice | Underlying knowledge sources, degree of fit with existing practices/values, clinical and patient experiences |
| Clinical encounter | Patient-provider interaction | Nature of the interaction between the patient and the provider | Individual, cognitive/affective, and communication factors that affect how the conversations is coordinated to achieve shared and personal goals |
| Recipients | Patient factors | Individuals who receive and are impacted by the innovation | Beliefs and preferences, biological or genetic conditions related to equitable implementation, health literacy, demographics, culturally relevant factors |
| Recipients | Provider factors | Clinicians, providers, and staff who administer the innovation | Knowledge and attitudes, competing demands, bias, demographics, culturally relevant factors |
| Recipients | Other recipients | Individuals and collective teams who influence access to the innovation | Resources, support, existing networks |
| Context | Local level | Inner context; clinic, department, ward, unit | Formal and informal leadership support, culture, previous experiences related to innovation |
| Context | Organizational level | Inner context; hospital or network | Organizational priorities, senior leadership or management support, learning networks |
| Context | Healthcare system | Outer context; broader resources, culture, leadership, orientation to learning | Leadership support, formal policies, structure of a system including location, resources, and complexity of practice |
| Societal influence | Economics | Structure of city, state, and country related to wealth and resources and what is exchanged for healthcare delivery | Healthcare coverage, insurance or private pay and how that influences care |
| Societal influence | Physical structures | Built environment | Physical ways and navigation to get to the innovation |
| Societal influence | Sociopolitical forces | Policies and procedures (formal or informal) in national and local governments that systematically inhibit or promote equitable health | Cultural norms, federal policies or laws |
| Social determinant of health | Economic stability | Poverty, employment status, ability to work | Disabilities, injuries, limiting conditions, childcare options |
| Social determinant of health | Education access and quality | Access to high-quality educational opportunities free from social discrimination that may impact health | Discrimination or bias that impacts disease problems or benefits |
| Social determinant of health | Health care access and quality | May include health insurance coverage, usual care management provider, coverage, distance/access to providers, communication with providers | Clinical or self referral, access to and coverage by insurance, telehealth options, wait times, language barriers to understanding care, access to electronic health records |
| Social determinant of health | Neighborhood and built environment | Presence of health or safety risks in places where people are born, live, learn, work, play, worship, and age | Hazardous or unhealthy sites, accessibility for disabled patients, access to transportation |
| Social determinant of health | Social and community context | Access to social support | Safety of neighborhoods, presence of caregivers, access to positive social environments such as patient support groups, health literacy |
| Other | Feedback/suggestions | Ideas presented by the participant regarding the project | Suggestion to talk with an individual or group |
| Other | Exemplar quotes | Quotes from participant that illustrates a theme or concept | Direct quote from the transcripts that capture a theme |
| Other | Emerging theme | Prominent ideas mentioned by participants that do not fall into another predetermined theme/code | Insights derived from the words or experiences of participants |
| Clinician demographics | Demographics | Clinician demographics, if provided | - |
| Clinician demographics | Institution | Institution of practice | - |
| Clinician demographics | Practice years | Years identified as working in specialty | - |
| Clinician demographics | Subspecialty | Area of clinical focus | - |
| Clinician demographics | Conditions treated | Types of cardiovascular genetic disorders the participant diagnoses or treats | - |

## Appendix III – HEIF aligned themes, examples, social determinants of health mappings, and exemplar quotes

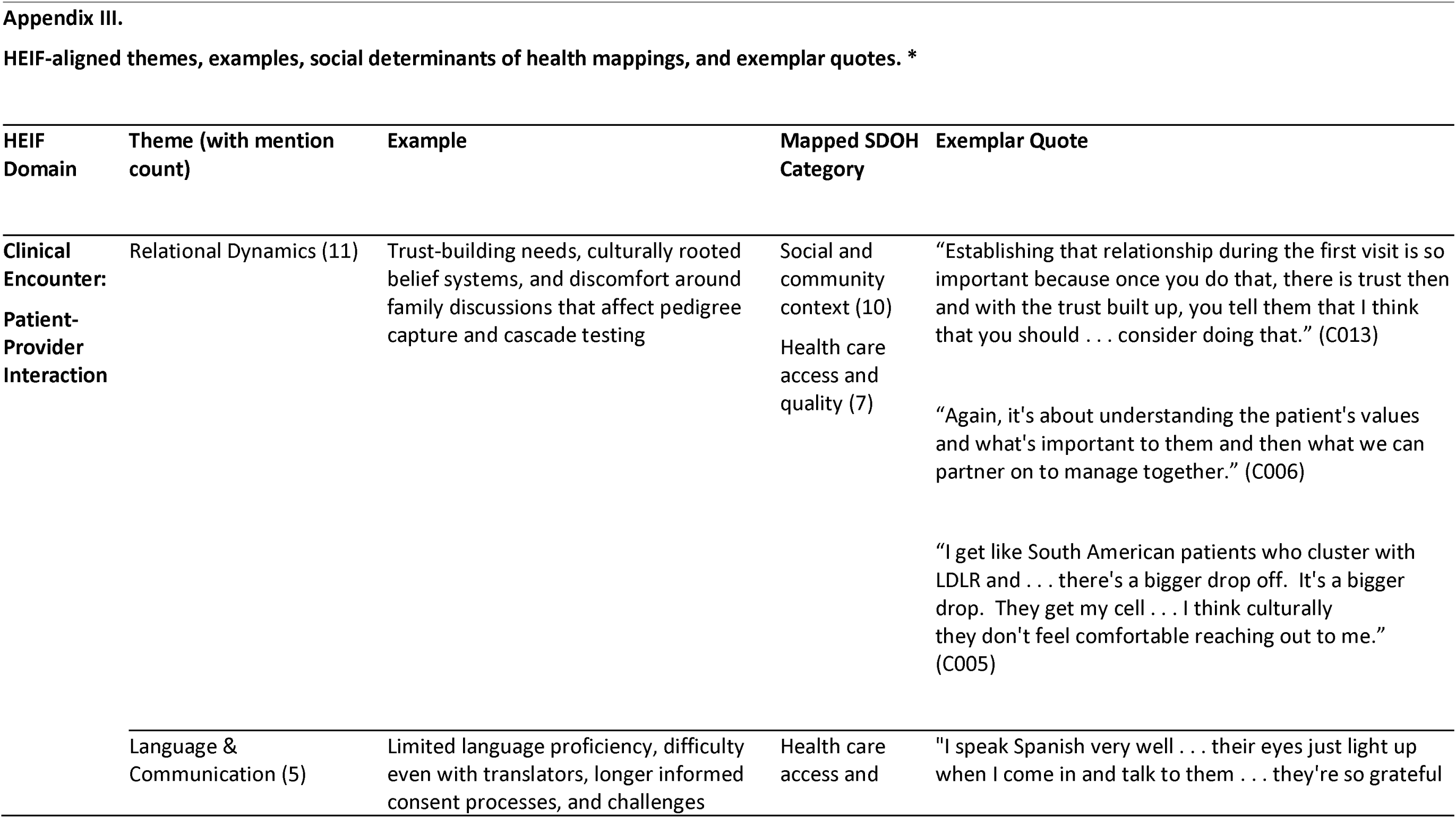
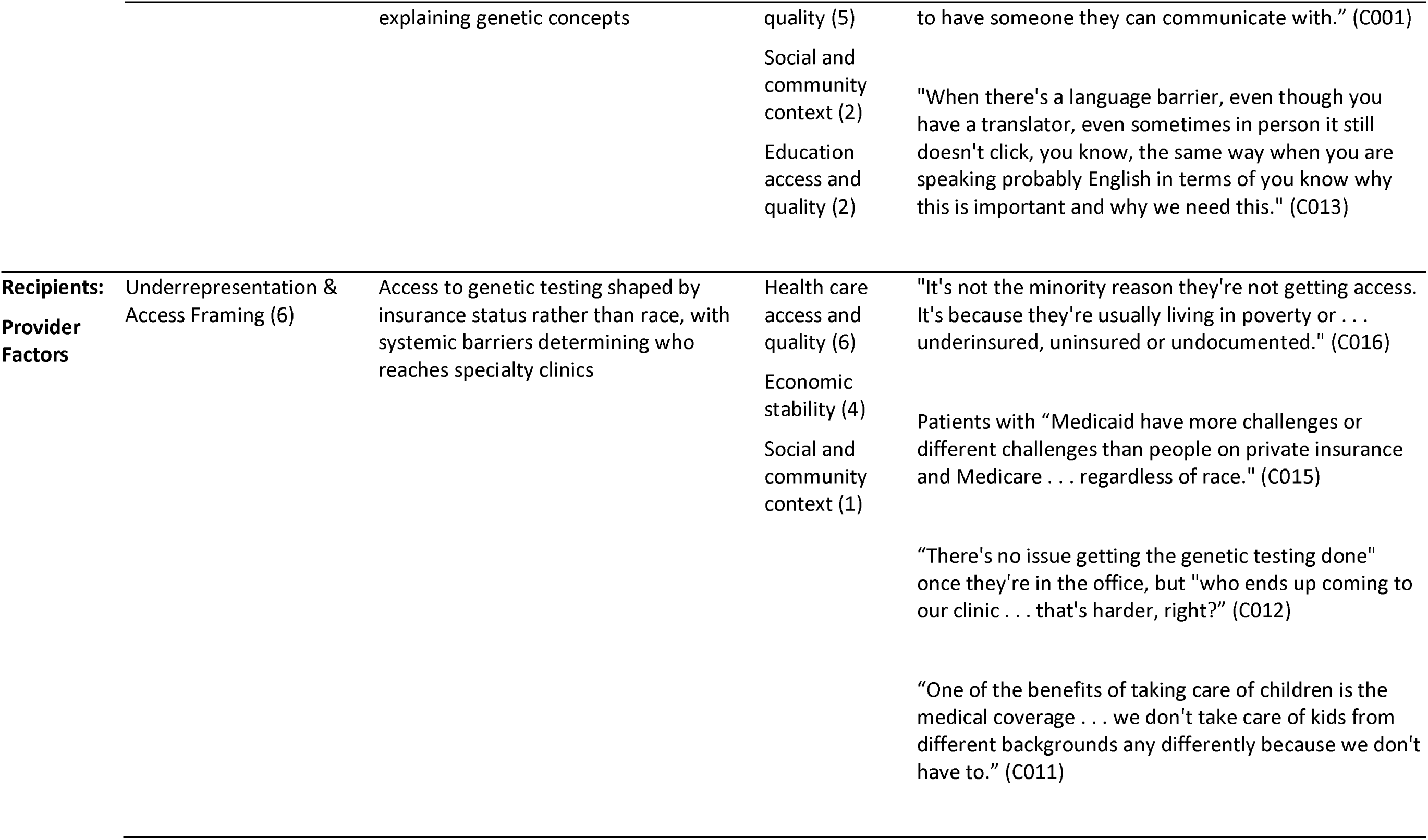
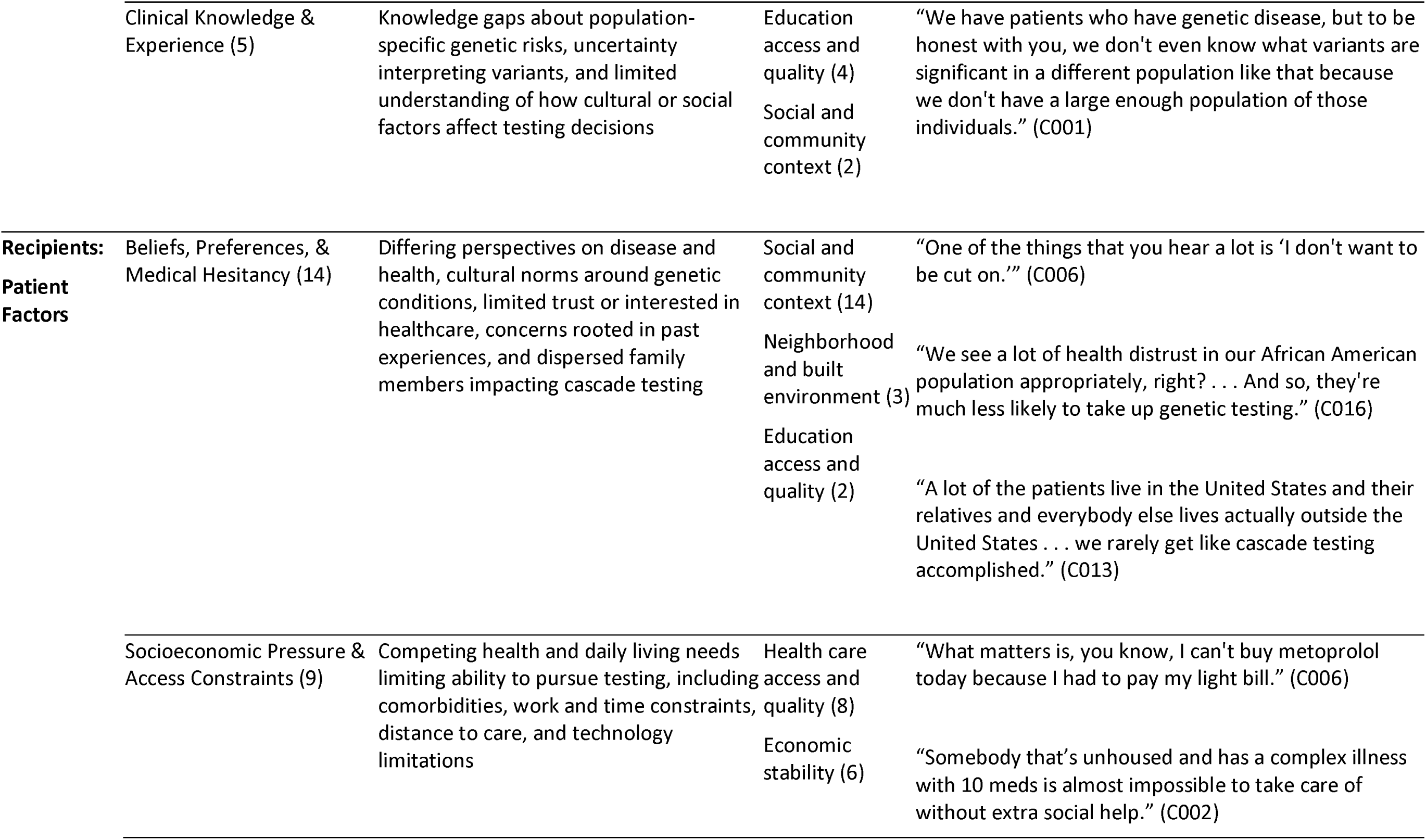
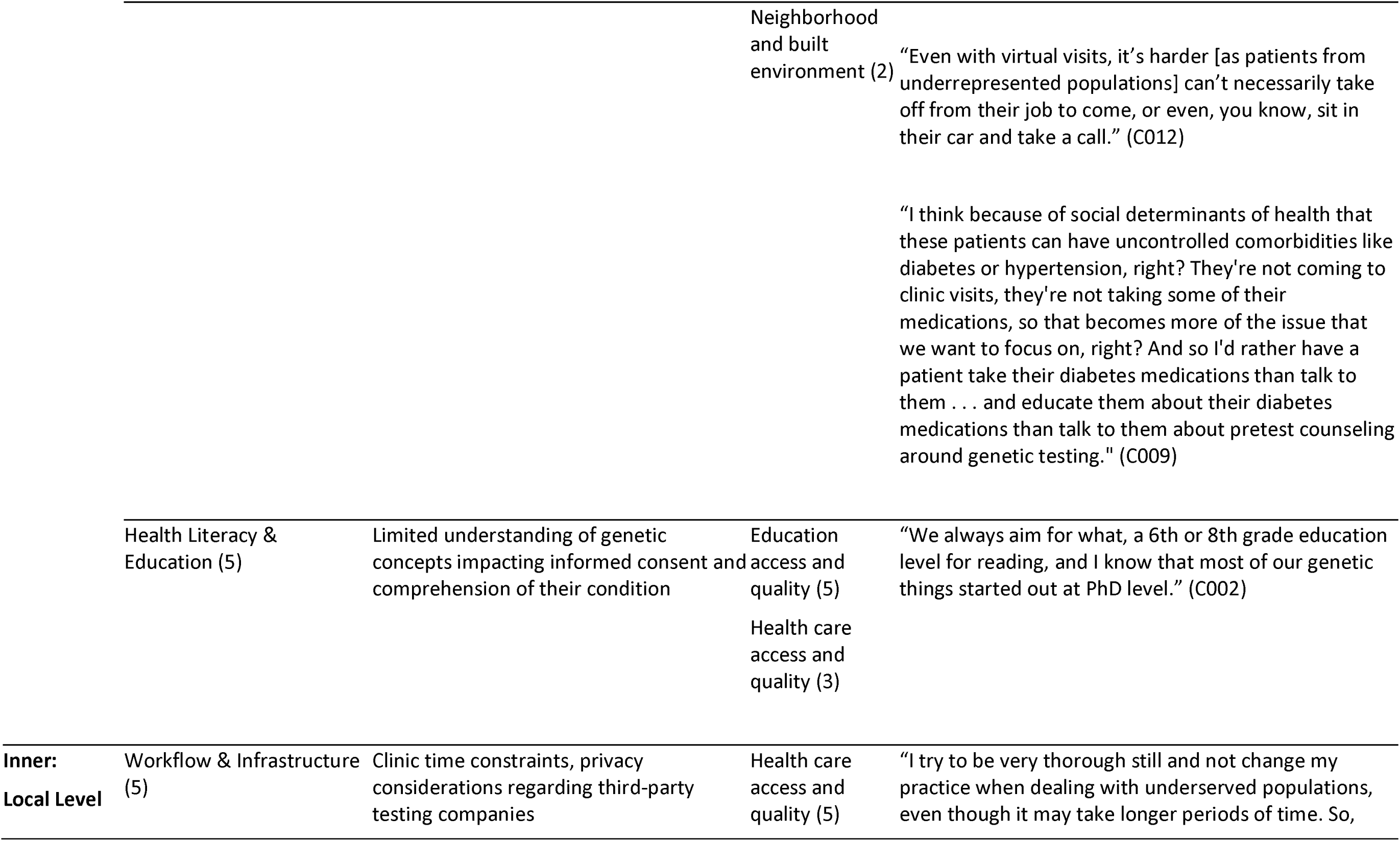
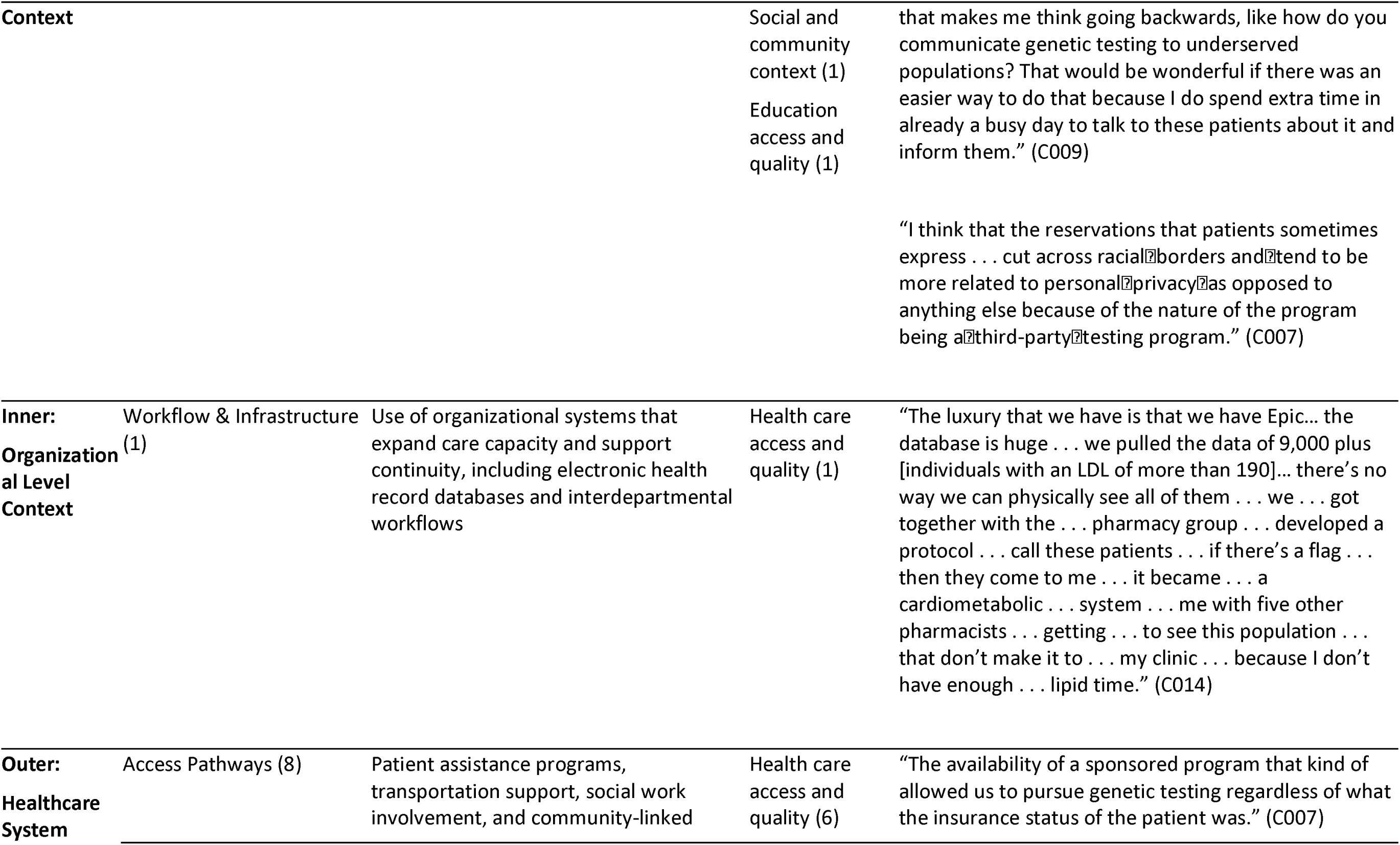
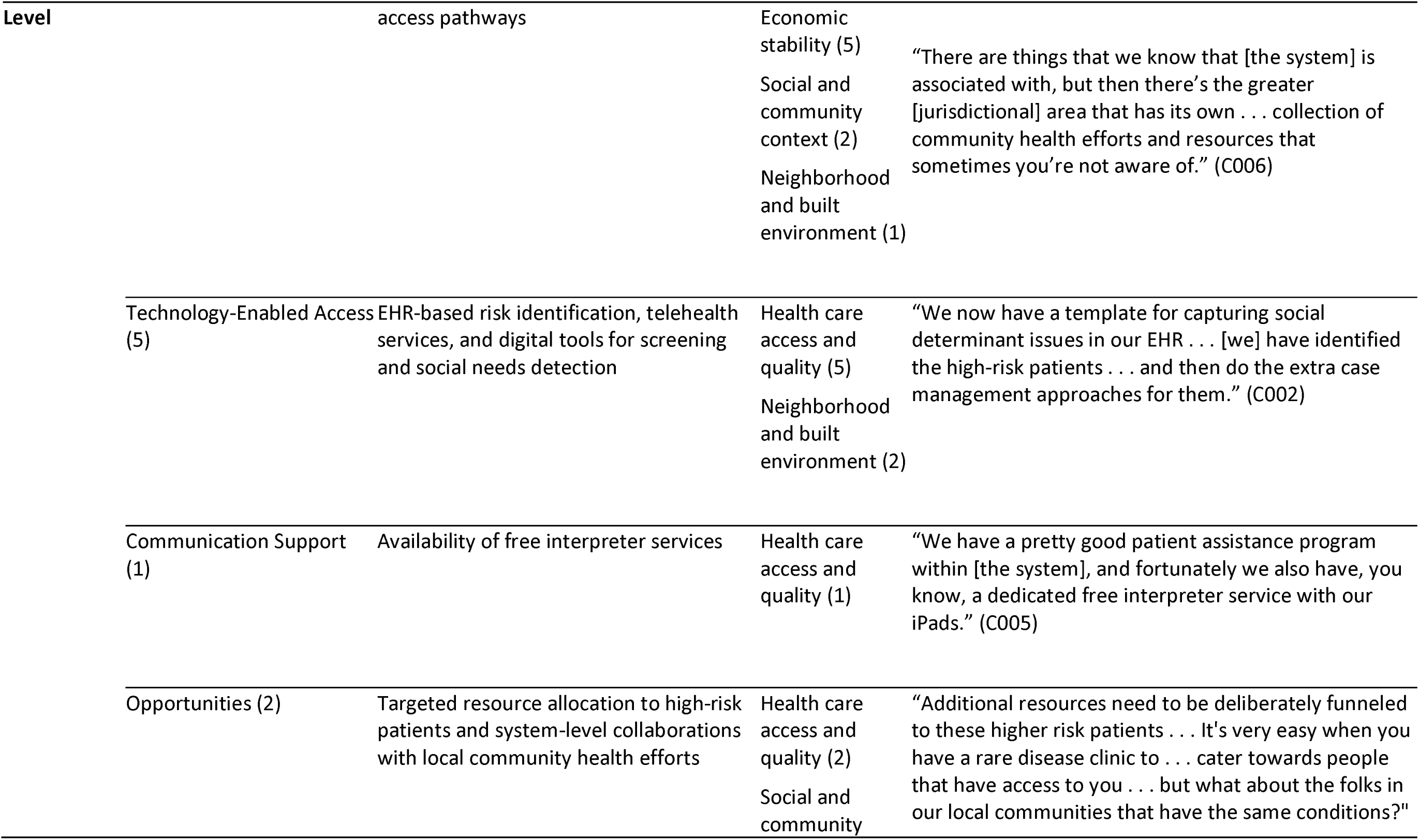
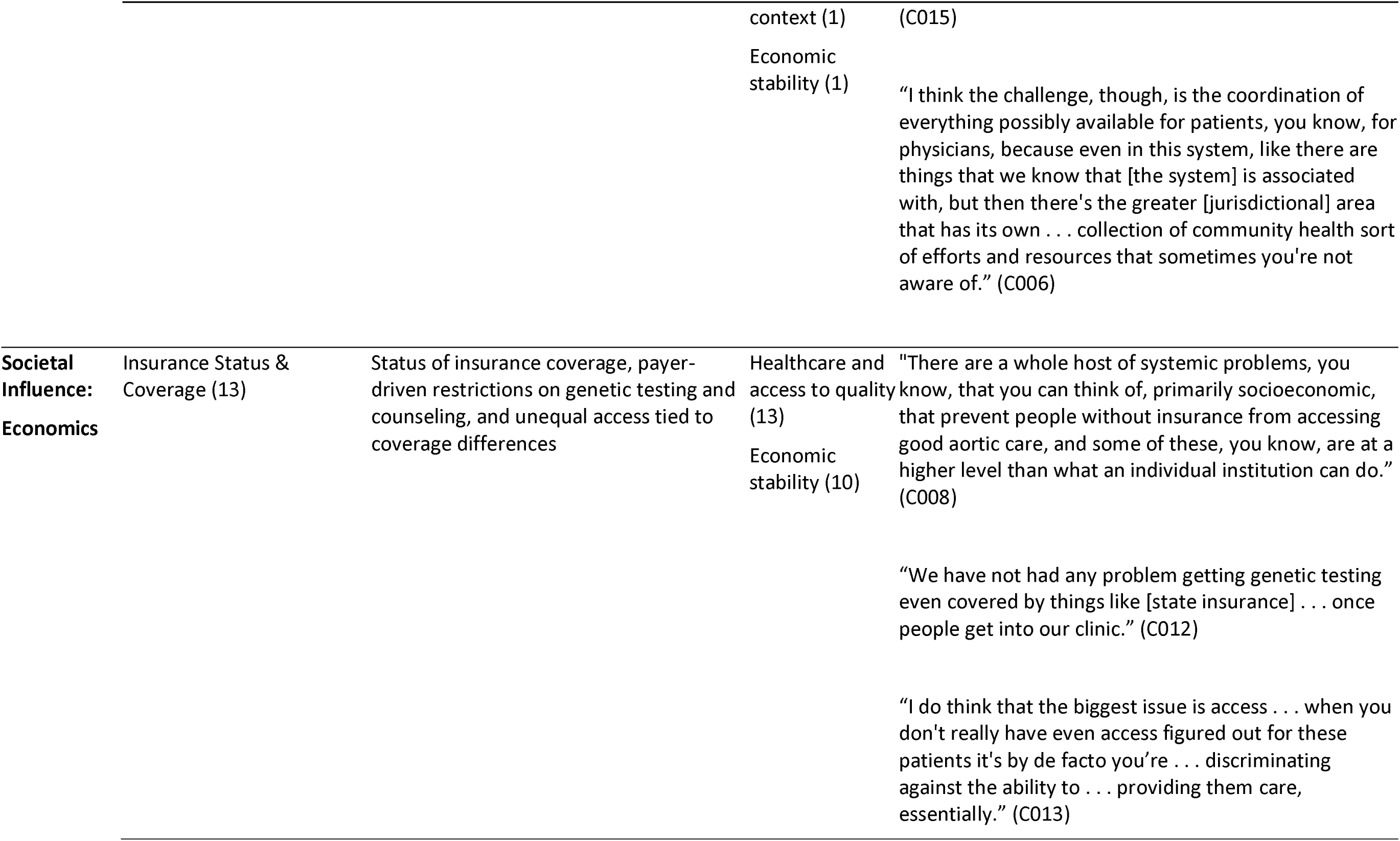
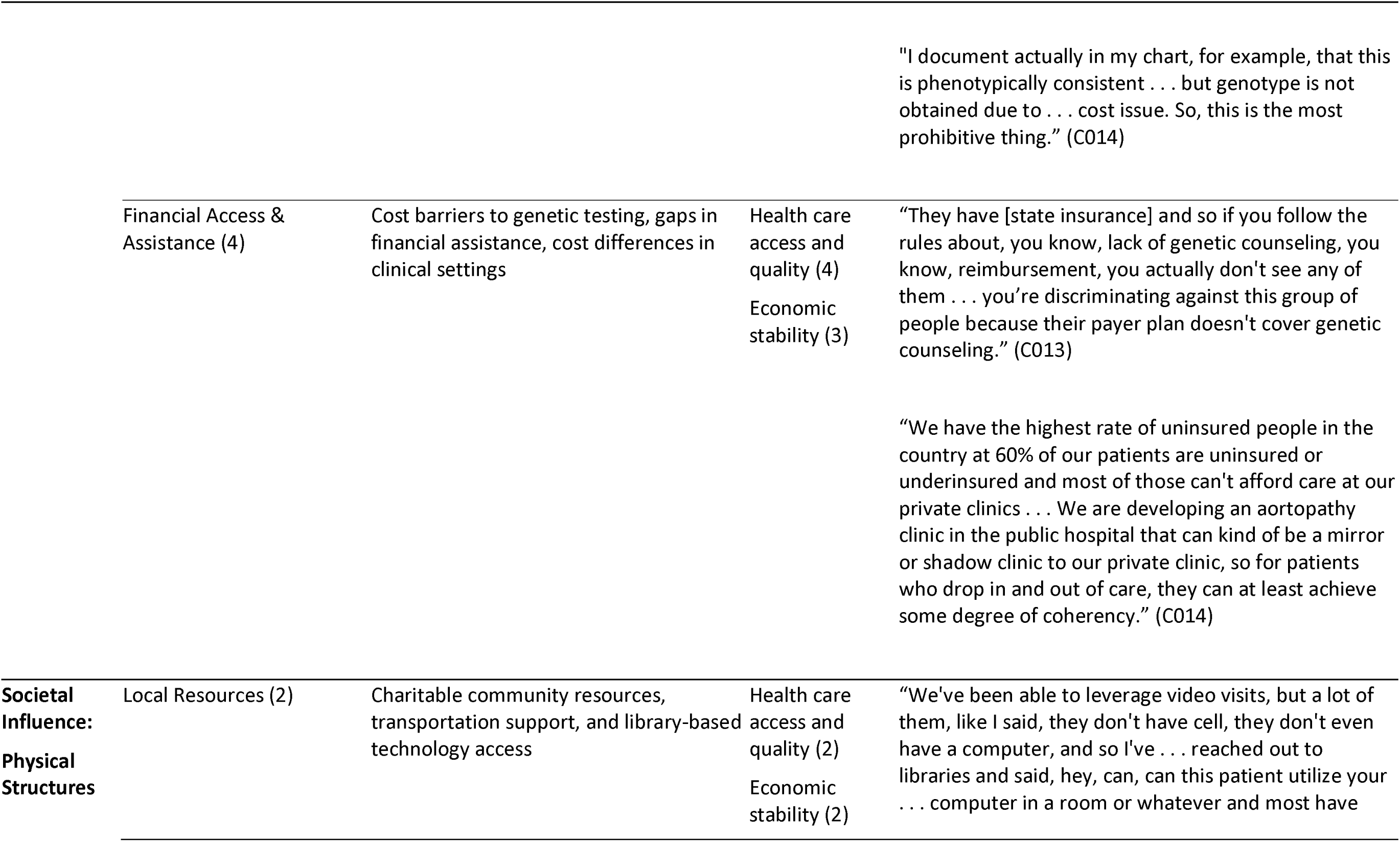
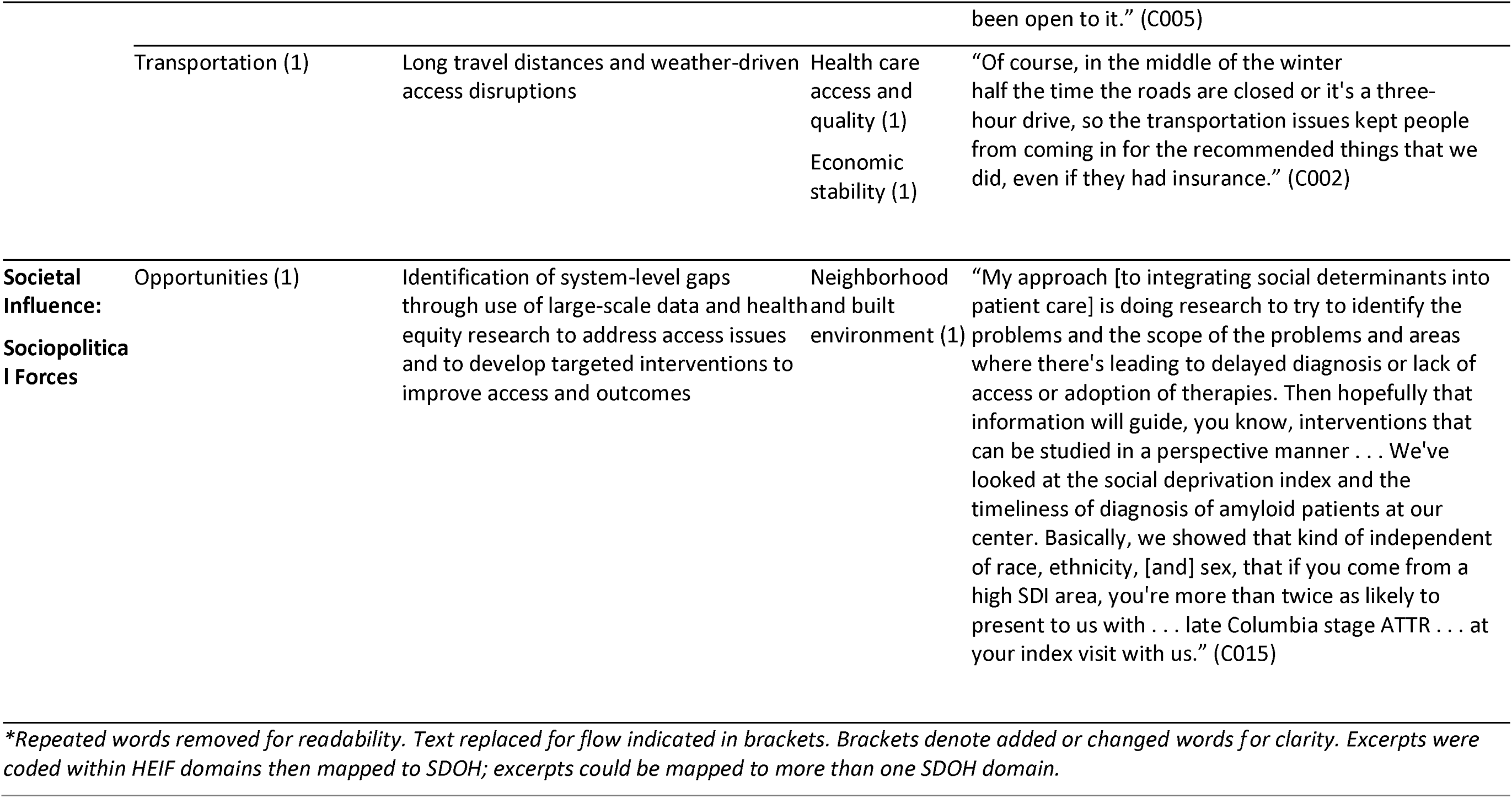

## References

1. World Health Organization: Cardiovascular diseases (CVDs). 2025. Available from https://www.who.int/news-room/fact-sheets/detail/cardiovascular-diseases-(cvds)

2. Chong B, Jayabaskaran J, Jauhari S, Chan S, Goh R, Kueh M, et al. Global burden of cardiovascular diseases: projections from 2025 to 2050. European Journal of Preventive Cardiology 2024;32(11):1001–1015.

3. Tada H, Fujino N, Hayashi K, Kawashiri M, Takamura M. Human genetics and its impact on cardiovascular disease. Journal of Cardiology 2022;79(2):233–239.

4. Jaffe K, Greene AK, Chen L, Ryan KA, Krenz C, Roberts JS, et al: Genetic researchers’ use of and interest in research with diverse ancestral groups. JAMA Network Open 2024;7(4).

5. Bains M, Qureshi N, Bajwa RK, Leonardi-Bee J, Hassanein ZM, Hassan S, et al: Ethnic inequities in genomic and precision medicine. NHS Race & Health Observatory 2024.

6. Geneviève LD, Martani A, Shaw D, Elger BS, Wangmo T: Structural racism in precision medicine: leaving no one behind. BMC Medical Ethics 2020;21(1).

7. Mudd-Martin G, Cirino AL, Barcelona V, Fox K, Hudson M, Sun YV, et al: Considerations for cardiovascular genetic and genomic research with marginalized racial and ethnic groups and Indigenous peoples: a scientific statement from the American Heart Association. Circulation: Genomic and Precision Medicine 2021;14(4).

8. Havranek EP, Mujahid MS, Barr DA, Blair IV, Cohen MS, Cruz-Flores S, et al. Social determinants of risk and outcomes for cardiovascular disease. Circulation 2015;132(9):873–898.

9. United Nations: The 17 goals | sustainable development. 2015. Available from https://sdgs.un.org/goals

10. Office of Disease Prevention and Health Promotion: Healthy People 2030. 2020. Available from https://odphp.health.gov/healthypeople

11. Suglia SF, Hidalgo B, Baccarelli AA, Cardenas A, Damrauer S, Johnson A, et al: Improving cardiovascular health through the consideration of social factors in genetics and genomics research: a scientific statement from the American Heart Association. Circulation: Cardiovascular Quality and Outcomes 2025;18(5):e000138.

12. Romagnoli KM, Salvati ZM, Johnson DK, Ramey HM, Chang AR, Williams MS: Genomics in nephrology: identifying informatics opportunities to improve diagnosis of genetic kidney disorders using a human-centered design approach. Journal of the American Medical Informatics Association 2024;31(6):1247–1257.

13. Woodward EN, Matthieu MM, Uchendu US: The health equity implementation framework: proposal and preliminary study of hepatitis C virus treatment. Implementation Science 2019;14.

14. AcademyHealth: Geisinger Health System. 2022. Available from https://academyhealth.org/

15. Motairek I, Chen Z, Makhlouf M, Deo S, Salerno P, Mentias A, et al: Mapping geographic proximity to cardiologists across the United States. Circulation: Cardiovascular Quality and Outcomes 2023;10.

16. Patton MQ: Qualitative research and evaluation methods. Thousand Oaks, Sage Publications; 2015.

17. Palinkas LA, Horwitz SM, Green CA, Wisdom JP, Duan N, Hoagwood K: Purposeful sampling for qualitative data collection and analysis in mixed method implementation research. Administration and Policy in Mental Health and Mental Health Services Research 2015;42:533–544.

18. Saunders B, Sim J, Kingstone T, Baker S, Waterfield J, Bartlam B, et al: Saturation in qualitative research: exploring its conceptualization and operationalization. Quality & Quantity 2018;52(4):1893–1907.

19. Ritchie J, Spencer L: Qualitative data analysis for applied policy research. In: Huberman AM, Miles MB, editors. The qualitative researcher’s companion. Thousand Oaks, Sage Publications: p. 305–329.

20. Gale NK, Heath G, Cameron E, Rashid S, Redwood S: Using the framework method for the analysis of qualitative data in multi-disciplinary health research. BMC Medical Research Methodology 2013;13(1):117.

21. O’Brien BC, Harris IB, Beckman TJ, Reed DA, Cook DA: Standards for reporting qualitative research: a synthesis of recommendations. Academic Medicine 2014;89(9):1245–1251.

22. O’Brien BC, Harris IB, Beckman TJ: The SRQR reporting checklist. EQUATOR Network 2025.

23. Lopez Santibanez Jacome L, Dellefave-Castillo LM, Wicklund CA, Scherr CL, Duquette D, Webster G, et al: Practitioners’ confidence and desires for education in cardiovascular and sudden cardiac death genetics. Journal of the American Heart Association 2022;7.

24. Musunuru K, Hershberger RE, Day SM, Klinedinst NJ, Landstrom AP, Parikh VN, et al: Genetic testing for inherited cardiovascular diseases: a scientific statement from the American Heart Association. Circulation: Genomic and Precision Medicine 2020;13(4).

25. Reza N, Alford RL, Belmont JW, Marston N: The expansion of genetic testing in cardiovascular medicine: preparing the cardiology community for the changing landscape. Current Cardiology Reports 2024;26(3):135–146.

26. Scherr CL, Kalke K, Ramesh S, Fakhari H, Dellefave-Castillo LM, Smith ME, et al: Integrating clinical genetics in cardiology: current practices and recommendations for education. Genetics in Medicine 2022;24(5):1054–1061.

27. Hundal K, Scherr CL, Fakhari H, Ramesh S, Dellefave-Castillo L, Duquette D, et al: Mapping the use of cardiovascular genetic services in pediatric clinical care: challenges and opportunities for improvement. Frontiers in Genetics 2025;15.

28. Borrell LN, Elhawary JR, Fuentes-Afflick E, Witonsky J, Bhakta N, Wu A, et al: Race and genetic ancestry in medicine—A time for reckoning with racism. N Engl J Med 2022;384(5).

29. Powell-Wiley TM, Baumer Y, Baah FO, Baez AS, Farmer N, Mahlobo CT, et al: Social determinants of cardiovascular disease. Circulation Research 2022;130(5).

30. Zhang Z, Jackson SL, Thompson-Paul AM, Yin X, Merritt RK, Coronado F: Associations between health-related social needs and cardiovascular health among US adults. Journal of the American Heart Association 2024.

31. Myers VD, Gerhard GS, McNamara DM, Tomar D, Madesh M, Kaniper S, et al: Association of variants in BAG3 with cardiomyopathy outcomes in African American individuals. JAMA Cardiol 2018;3(10).

32. Morris AA, Masoudi FA, Abdullah AR, Banerjee A, Brewer LC, Commodore-Mensah Y, et al: 2024 ACC/AHA key data elements and definitions for social determinants of health in cardiology: a report of the American College of Cardiology/American Heart Association Joint Committee on Clinical Data Standards. Circulation: Cardiovascular Quality and Outcomes 2024;10.

